# Efficacy, Safety and Cost-effectiveness of Atorvastatin 40mg versus 80 mg in South Asian Patients with acute coronary syndrome: A protocol for randomised clinical trial

**DOI:** 10.1101/2024.08.09.24311739

**Authors:** Kavindya Fernando, Nilshan Fernando, Chiranthi Welhenge, Shashima Liyanage, B.K.T.P. Dayanath, Shamila De Silva, Chamila Mettananda

## Abstract

**Introduction:** Most guidelines recommend high-intensity statins for the secondary prevention of Acute Coronary Syndrome (ACS). However, it has been observed that South Asians are responsive to lower doses of statins to achieve the recommended target levels of low-density lipoprotein cholesterol (LDL-C). However, published data on this subject is limited. Therefore, we aim to compare the efficacy, safety and cost effectiveness of atorvastatin doses (80mg vs 40mg) in lowering the LDL-c levels to the less than 70 mg/dL within 12 weeks, among patients with incident ACS.

**Methods and analysis:** This single-centre, prospective, randomised, controlled, open-labelled clinical trial is being conducted among patients naïve for statins presenting with incident ACS to the Colombo North Teaching Hospital, Ragama, Sri Lanka. All participants are randomised to two groups, to receive oral atorvastatin 40 mg nocte, or 80 mg nocte. The percentage of patients achieving treatment goals, the percentage with statin induced adverse drug reactions and the mean cost to achieve treatment goals will be compared between the two groups in 6,12 and 24 weeks. Outcome will be analysed for the intention-to-treat population.

**Ethics and dissemination:** Ethical approval for this study has been obtained from the Ethics Review Committee of the Faculty of Medicine, University of Kelaniya (P/28/05/2022). This trial is registered in the Sri Lanka Clinical Trial Registry (SLCTR/2023/003). The results from this study will be disseminated as scientific publications in reputed journals.

**Key points of this study:** What is already known on this topic?

- While the European guideline recommendation is to initiate atorvastatin at 80 mg dose for ACS, it has been observed that South Asians respond well to lower dose. Additionally, South Asians tend to experience more adverse effects with higher doses of statins. However, reports on differential action of atorvastatin in South Asians is limited.

**What this study adds?:**

- This study explores the efficacy, safety, and cost-effectiveness of a lower dose of atorvastatin (40 mg daily) compared to guideline recommended dose of 80mg in a South Asian cohort.

**How might this study affect research, practice or policy?:**

- The findings may influence statin dose recommendation for South Asians/Sri Lankans which will help to reduce unnecessary side effects and be cost saving as well in secondary prevention of ACS.

## INTRODUCTION

Acute coronary syndromes (ACS) are the leading cause of death worldwide (1), with a rising prevalence in South Asia (2,3). Once ACS is diagnosed, patients must adhere to secondary prevention strategies to reduce mortality risk, with a key objective being the reduction of low-density lipoprotein cholesterol (LDL-C) levels to less than 70 mg/dL (1.8 mmol/L) (4–6). Statins are the first line therapy used to achieve this goal. In Sri Lanka, Atorvastatin is the most used statin in state hospitals.

While numerous studies have demonstrated the efficacy of atorvastatin at an 80 mg dose in achieving target LDL-C levels, Sri Lankan physicians often prescribe a 40 mg dose. This practice is based on observed clinical responses. This is also possibly due to differences in pharmacokinetics between South Asians and Western populations (7). However, there is no clinical evidence to support this practice.

Eighty milligrams of atorvastatin is a significantly high dose, and higher doses increase the likelihood of development of both dose-related and non-dose-related adverse effects. Given that Sri Lanka’s healthcare system provides free medication, including statins, reducing the standard dose from 80 mg to 40 mg could lower healthcare costs without compromising patient outcomes. This study aims to establish scientific evidence for the efficacy, safety, and cost-effectiveness of using 40 mg of atorvastatin, compared to 80 mg, as a secondary preventive measure for cardiovascular diseases in ACS patients.

Hence this study will provide clinical evidence for prescribing 40 mg of atorvastatin for patients with ACSs in clinical practice.

## METHOD AND ANALYSIS

This proposal was developed in accordance with the National Institute for Health and Care Excellence (NICE) guideline recommendations on lipid modification therapy for cardiovascular disease prevention and cholesterol clinical practice guideline by American Heart Association 2018 (4–6).

### Study design and setting

This study is an ongoing single centred, randomised, controlled, open labelled clinical trial, conducted in the University Medical Unit of Colombo North Teaching Hospital, Ragama, Sri Lanka. The study evaluates the efficacy, safety and cost-effectiveness of atorvastatin 40 mg compared to 80 mg as a secondary preventive measure in South Asian patients with ACS.

### Study hypothesis

In South Asian patients experiencing incident ACS, a daily dose of 40 mg of atorvastatin is adequate, safer and more cost-effective in achieving the target LDL-C levels to less than 70 mg/dL, compared to a dose of 80 mg.

### Study population and eligibility criteria

All consenting patients above 18 years of age with incident ACS admitted to the medical casualty ward of the University Medical Unit, Colombo North Teaching Hospital, Ragama will be included.

#### Inclusion criteria

- Male or female patient aged 18 years or older
- Patients presenting with incident ACS as defined by American Heart Association/ American College of Cardiology guideline in 2014 (8).
- Patients with ability to understand and follow study related instructions
- Patients providing written informed consent prior to any study-specific criteria

#### Exclusion criteria

- Patients with familial hypercholesterolemia (previously identified patients and patients screened using Simon Broome criteria (9))
- Patients diagnosed with type 2 diabetes mellitus at screening visit
- Patients diagnosed with chronic kidney disease (CKD) at screening visit
- Patients who are already on statin therapy
- Patients with known hypersensitivity to the study treatment
- Patients receiving concomitant treatment with cytochrome P450 3A4 (CYP3A4) inhibitors
- Pregnant or lactating mothers
- Any other condition or therapy which would make the patient unsuitable for this study or will not allow participation for the full planned study period (e.g. active malignancy or other condition limiting life expectancy to <12 months)

### Sample size

The sample size of this study was determined based on an anticipated response rate of 47% in the group receiving 40 mg atorvastatin nocte, as reported in a prior study in India (10). The calculated minimal sample size is 70 patients with 35 patients each in the intervention and control groups, considering type I error as 0.05 and a power as 80%. Accounting for a potential 10% dropout rate, the study aims to recruit a total of 78 patients.

### Subject enrolment and randomisation

Researchers will visit the University Medical Unit on ‘Day 0’ to identify patients presenting with incident ACS who fulfil the eligibility criteria. Each patient will be given a patient information sheet in their preferred language and will get the opportunity to clarify doubts with the investigators before providing consent. Informed written consent will then be obtained prior to enrolment into the study.

Upon recruitment, an interviewer-administered questionnaire will be completed, collecting data on socio-demographic background, past medical history, and family history. Additionally, anthropometric measurements, including height, weight, and waist circumference will be recorded.

Once a physician decides the need for “high-intensity” statin therapy, patients will be randomly assigned to either the intervention group or control group using a random number generator list. The intervention group will receive 40 mg of atorvastatin daily, while the control group will receive 80 mg of atorvastatin daily, which is the recommended dose according to NICE guidelines (Figure 1).

**Figure 1:**
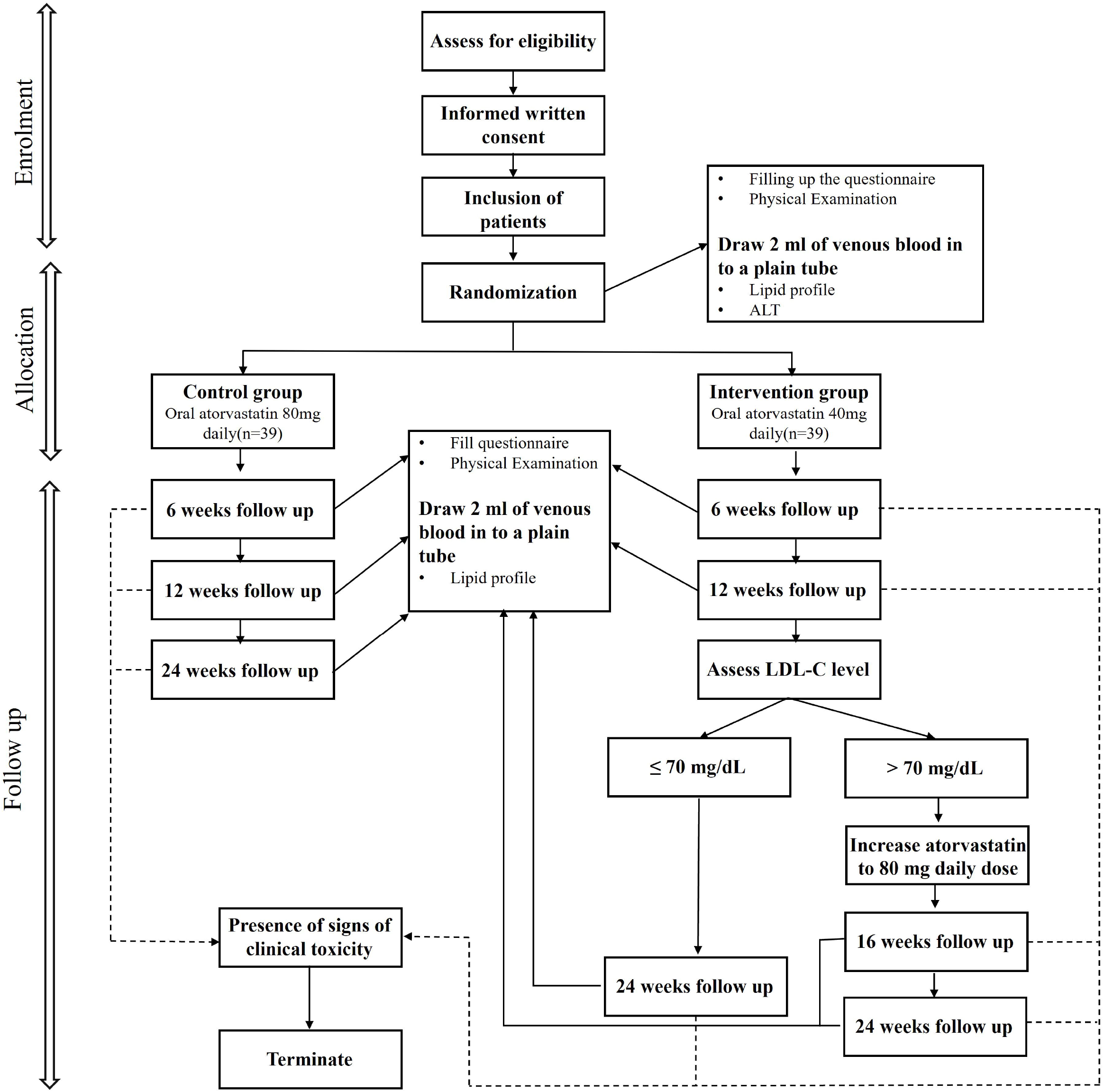
Study design of the project. Abbreviations: ALT – alanine transaminase; LDL-C – low-density lipoprotein cholesterol

### Intervention

Patients in the intervention group will receive oral atorvastatin at dose of 40 mg, while patients in the control group will receive atorvastatin at a daily dose of 80 mg. The atorvastatin formulations are manufactured by Gamma Interpharm (Pvt) Ltd, Sri Lanka. All other standard treatments and interventions that include percutaneous coronary interventions (PCI) and other cardiac interventions will be continued.

### Study procedure

#### Clinical evaluation

All patients will be reviewed at enrolment by a consultant physician to determine their suitability for high-intensity statin therapy. Following randomization and initiation of the assigned atorvastatin dose, these patients will be reviewed at the Medical Clinic of Colombo North Teaching Hospital at 6 weeks, 12 weeks and optionally 16 weeks, with a final review at 24 weeks (Figure 1). During each review visit, a trained medical officer will complete the questionnaire to assess drug adherence, current medical status and any previously unknown adverse effects associated with atorvastatin. A comprehensive physical examination will be performed to measure weight and waist circumference, as well as to evaluate tolerance and to detect any adverse effects of the medication.

#### Laboratory evaluation

At enrolment, a baseline venous blood sample will be drawn from each patient to evaluate the lipid profile, including total cholesterol, high-density lipoprotein cholesterol (HDL-C), LDL-C and triglycerides, and alanine transaminase (ALT) levels. Subsequent lipid profiles will be measured at each clinic visits at 6 weeks, 12 weeks, 16 weeks and 24 weeks. In the event of any adverse effects, ALT and creatine phosphokinase (CPK) levels will be assessed as needed.

Lipid profiles will be performed under 12 hour fasting conditions. The first sample at enrolment will be drawn within 24 hours of onset of chest pain, as recommended (11). Total cholesterol, HDL-C and triglycerides will be analysed using enzymatic colorimetric methods using Mindray BA-88A semi-automated chemistry analyser (Shenzhen Mindray Bio-Medical Electronics Co. Ltd, China). Internal quality control will be maintained at two levels: Level 1 for physiological range and Level 2 for pathological range, with EXATROL quality control materials supplied by BIOLABO SAS, France. The laboratory will participate in the National External Quality Assurance Scheme (NEQAS) of Sri Lanka, administered by the Medical Research Institute, Colombo. LDL-c will be calculated using Friedewald equation (12) and non-HDL-C will be determined by subtracting HDL-C from total cholesterol.

#### Safety evaluation

During follow up visits at 6 weeks, 12 weeks 16 weeks (optional), and 24 weeks, patients will be interviewed by a trained medical officer to monitor for both known and unknown adverse effects associated with atorvastatin treatment. The potential adverse effects include hepatitis, myositis, myalgia, and gastric discomfort. If any signs of toxicity are observed, patients will be admitted to the University Medical Unit, Colombo North Teaching Hospital, Sri Lanka, for further management and monitoring, and ALT and CPK levels will be assessed. Such patients will be discontinued from the study.

All participants will be provided with contact information of the investigators to address any questions regarding the trial and to report any suspected adverse effects throughout the study.

#### Cost effectiveness evaluation

The total amount of money spent for medication will be calculated separately for both arms. The mean cost spent on medication to achieve the targeted LDL-C level will be compared between two groups. The “effectiveness” will be measured from health benefit achieved (number of patients who reached target LDL-C reduction within 12 weeks). The average cost effectiveness ratio and incremental cost effectiveness ratio will be calculated.

### Outcome measures

#### Primary outcome

Reduction of LDL-C levels to less than 70mg/dL at 12 weeks will be the primary outcome.

#### Secondary outcomes

1. Percentage reduction of LDL-C from baseline
2. Non-HDL-C reduction of at 12 weeks and 24 weeks
3. Safety profile and adverse effects of treatment
4. Cost-effectiveness of the treatment

### Statistical analysis

Data will be analysed using univariate and multivariate analysis by SPSS version 26. The analysis will follow an intention-to-treat approach, including dropouts and patients who discontinue the study. Non-parametric tests will be used to compare the adverse events in both arms. Statistical tests will be conducted at a 5% two-sided significance level. Comparative outcomes will be summarized with 95% confidence intervals and reported in accordance with the CONSORT guidelines.

### Data management and monitoring

All collected data will be anonymized and kept confidential. Questionnaires and laboratory reports will be securely stored in locked cabinets, accessible only to the investigators. Personal identification details will remain confidential and will not be disclosed. Blood samples will be securely destroyed after analysis and will not be used for any additional testing. The electronic database will be maintained as a password-protected file. All information will be disseminated solely through publication.

### Ethical considerations

All patient management decisions will be made by the clinical management team of the University Medical Unit in accordance with the Unit’s protocols. Investigators will be responsible solely for administering the assigned dose of atorvastatin (either 40 mg or 80 mg). Patients who experience severe adverse events, whether related or unrelated to the treatment, or those who cannot tolerate atorvastatin, will be discontinued from the study. Suspected adverse events will be reported according to the national guidelines for adverse event reporting. Reports will be submitted in the specified format to the Director of Medical Technology and Supplies at the National Medicines Regulatory Authority of Sri Lanka, and to the Adverse Drug Reaction Monitoring Unit of the Department of Pharmacology, Faculty of Medicine, University of Colombo. Participants have the right to withdraw from the study at any time without needing to provide a reason.

Ethical approval for the study has been granted by the Ethics Committee of the Faculty of Medicine, University of Kelaniya, Sri Lanka (P/28/05/2022). The trial has also been registered with the Sri Lanka Clinical Trial Registry (SLCTR/2023/003).

### Termination of the trial

The trial will be terminated if

- The planned sample size is recruited
- New information or other evaluation regarding the safety or efficacy of atorvastatin dose that indicates a change in the known risk/benefit profile, such that the risk/benefit is no longer acceptable for subjects participating in the trial.
- Significant violation of good clinical practice (GCP) that compromises the ability to achieve study objectives or compromises subject safety, occurs

### Study status

The trial commenced in May 2023 according to the Protocol Version 2.0, dated August 15, 2022. Recruitment is currently ongoing, and to date, 48 patients have been enrolled in the study.

### Patient and public involvement

The research question for this clinical trial was formulated based on clinical practices observed among Sri Lankan physicians and cardiologists, who prescribe 40 mg of atorvastatin to patients following acute coronary syndromes, noting satisfactory responses. Reports generated from the trial will be accessible to all participants and utilized in their standard management as needed. The study results will be shared with participants and integrated into standard clinical practices.

## DISCUSSION

The prevalence of ACS is rising in South Asia, creating a significant challenge to public health. Effective secondary prevention strategies are crucial for reducing recurrent events and enhancing patients’ quality of life. In Sri Lankan medical practice, it has been observed that patients with ACS often respond well to a daily night dose of atorvastatin 40 mg. Despite this, there is a lack of evidence supporting this atorvastatin dose. Consequently, Sri Lankan physicians continue to prescribe 40 mg, although current guidelines recommend 80 mg based on data primarily from Western studies (13).

Studies in the Western world have predominantly used atorvastatin at 80 mg, which is supported by clinical guidelines as the maximum tolerated dose. A study conducted in Singapore suggests that Asian patients require similar statin doses to achieve target cholesterol levels as their Caucasian counterparts (14). However, evidence from India shows varying results. Agrawal et al. (2018) reported mean percentage reductions in LDL-C of 47.18 ± 20.81% and 50.03 ± 18.06% at 3 months for atorvastatin 40 mg and 80 mg, respectively, in a population with dyslipidaemia. But studies, among ACS group were not found. However, there are no reported Sri Lankan studies (15).

Most research work on lipid-lowering agents has been conducted among Caucasians, with limited data from Asian populations. It is well-established that therapeutic agents can have different effects in South Asians compared to Caucasians (7). Observations suggest that South Asian patients may achieve similar therapeutic effects with lower doses of statins, potentially due to higher plasma levels of statins in this population (16,17). Pharmacokinetic studies have noted higher plasma concentrations of statins in Asians without indicating pharmacokinetic safety issues at equivalent doses (18). Additionally, Ranasinghe et al. (2023) identified that certain minor allele frequencies might increase the risk of statin-induced myotoxicity in Sri Lankans (7).

Given these factors, it is essential to determine the appropriate statin dosage for Sri Lankan individuals to optimize therapeutic outcomes while minimizing adverse effects.

## Data Availability

All data produced in the present study are available upon reasonable request to the authors

## LIMITATIONS

One limitation of this study is the lack of prior research specifically addressing this topic. However, given that it is well-observed that the Sri Lankan population responds adequately to a 40 mg dose of atorvastatin, the findings will be valuable for researchers planning similar investigations and applying the results to their studies. Although an Indian study on a related topic (the CURE trial) exists it was not conducted among patients with ACS. Additionally, the study’s single-centre design may limit the generalizability of the findings.

